# Shortening heart rate variability measurement time to 1 minute using deep learning: Implication of real-time measurement of heart rate variability

**DOI:** 10.1101/2023.10.23.23297390

**Authors:** Junghwa Shin, Byung-Chae Lee, Kee Sam Jeong, Kwang Yoon Kim, Bom-Taeck Kim

## Abstract

Heart rate variability (HRV) is an effective predictor of cardiovascular diseases. The current standard 5-min recording time is lengthy compared with routine clinical examinations such as blood pressure measurement. Previous studies have observed that the indices of 3-min HRV data are as clinically meaningful as those of 5-min HRV data; however, shorter durations are considered unreliable, and there have been no attempts to challenge this notion. This study aimed to validate the outcomes of 1-min HRV recordings reconstructed using deep learning algorithms. Three-minute HRV recordings from 34,885 participants were included in the analysis. Of the recordings, 60% (20,931), 30% (10,465), and 10% (3,489) were allocated to the training, validation, and test sets, respectively. Data from 1-min excerpts of the 3-min recordings were used as the input for the deep learning models to predict the data of the 3-min recordings. Various deep learning models were applied to each indicator, and the model that produced the lowest mean absolute error was selected as that particular indicator’s learning model. There was no statistical difference between the values of the 1-min recordings reconstructed by deep learning and those of the 3-min recordings. The 1-min recordings reconstructed by deep learning demonstrated a higher correlation with the 3-min recordings when compared with the 1-min recordings that were not processed by deep learning. They also strongly agreed with the 3-min recordings in the Bland–Altman analysis. The 1-min HRV recordings reconstructed by deep learning were as reliable as the 3-min HRV recordings, suggesting that a 1-min recording could serve as a proxy for real-time HRV monitoring in the future.

## Introduction

The heart rate variability (HRV) test is a simple and effective noninvasive tool that can predict cardiovascular events by assessing the autonomic nervous system function [1]. The interaction between the heart and brain is affected by rhythms generated by higher frequency changes (e.g., vagal outflow, blood pressure and respiratory control) and slower modulations (e.g., circadian rhythms, thermoregulation, and hormonal regulation) [2,3]. The heart rate tachogram, a plot of a time sequence of R-R intervals is the most common form for observing these changes [3]. Heart rate fluctuations within specific time intervals yield time, frequency, and nonlinear indices. The ECG recording time of a conventional short-term HRV includes at least 10 cycles of the lower-frequency bound of the investigated component, for instance, 1-min for high-frequency (HF) (0.15–0.4 Hz), 2 min for low-frequency (LF) (0.04–0.15 Hz), and 5-min for very low-frequency (VLF) (≤ 0.04 Hz) [4]. An ECG recording time of longer than 5-min provides an acceptable resolution that distinguishes different frequency domains using fast Fourier transform (FFT), based on which the 5-min recording time became standard in HRV testing [5]. However, numerous variations in HRV recording times are used in different clinical and nonclinical settings and range from < 5-min (ultrashort-term), to 5-min (short-term), and ≥ 24 h (long-term) [4].

Developments in mobile and wearable devices have increased the demand for ultrashort-term HRV to assess fitness and overall health in real time. However, the greatest advantage of ultrashort-term HRV is the potential to save lives because it quickly predicts cardiac mortality and malignant arrhythmias [2]. Ultrashort-term HRV testing may also contribute to a range of situations where real-time assessment of autonomic functions is required, for instance, athletes in training, workers in highly demanding jobs, and critically ill patients in ICUs. The increasing need for, and benefits of, ultrashort-term HRV tests are evident, yet questions arise about the precision of the data obtained within the shortest HRV test time possible [6].

Previous studies showed that some indices such as mean heart rate (mHR), the square root of the mean of the sum of the squares of differences between adjacent NN intervals (RMSSD), HF from ultrashort-term HRV could reliably estimate short-term HRV [7–9]. The time-sensitive indices such as standard deviation of all NN intervals (SDNN), total power (TP), and VLF required at least 4-min (240 seconds) [8]. Measurements from ultrashort-term HRV tests have thus far been considered unreliable; however, no study has tried to challenge this time barrier using deep learning.

This study attempted to validate 1-min HRV data reconstructed by deep learning models by comparing them with those of the 3-min data to establish their reliability and procedural feasibility.

## 2. Materials and Methods

### 2.1 Materials

43,504 HRV recordings were randomly selected from six different medical centers that used the same HRV testing device (SA3000P; Medicore, Seoul, Korea). The recordings were from patients who visited for regular health checkups between January 2011 and December 2019. Demographic information, such as age, sex, medical, and drug history, was not considered in the analysis; however, age and sex distribution were even in the 3-min and 1-min groups. At the time of the test, patients were either seated or lying down. Patients were previously informed to avoid confounding factors, such as caffeine or food intake. The time of day was not specified; however, most tests were conducted in the morning.

8,619 recordings showing bradycardia, tachycardia, arrhythmias, or artifacts were excluded. In total, 34,885 HRV recordings were included in the analysis. Data from the original 3-min recording (Raw-3) served as the target. From Raw-3, a 1-min recording (Raw-1) was selected 30 s after the start of the test. Raw-1 was processed using deep learning models to produce outcome (Rec-1).

### 2.2 Learning label

The indices used in this study were the mHR, RMSSD, SDNN, TP, power in the VLF range, power in the LF range, and power in the HF range (Table 1). The mean RR interval (mRR), LF power in normalized units (LFn), and HF power in normalized units (HFn) were excluded from the learning process because these indices can be directly calculated from mHR, LF, and HF.

**Table 1.** HRV indices.

| Indices | Unit | Explanation |
| --- | --- | --- |
| mRR | ms | mean R-R interval |
| mHR | bpm | mean Heart Rate average heart rate |
| SDNN | ms | Standard deviation of all NN intervals |
| RMSSD | ms | The square root of the mean of the sum of the squares of differences between adjacent NN intervals |
| TP | ms<sup>2</sup> | Total Power |
| VLF | ms<sup>2</sup> | Power in the very low-frequency range $\leq 0.04$ Hz |
| LF | ms<sup>2</sup> | Power in the low-frequency range 0.04–0.15 Hz |
| HF | ms<sup>2</sup> | Power in the high-frequency range 0.15–0.4 Hz |
| LFn | n.u. | LF power in normalized units $LF / (Total\ Power - VLF) \times 100$ |
| HFn | n.u. | HF power in normalized units $HF / (Total\ Power - VLF) \times 100$ |
| LF/HF |  | Ratio LF/HF |

### 2.3 Input data for the deep learning models

Generally, the training data for the deep learning models should be uniform in size; however, the number of heartbeats in Raw-1 varied according to each individual’s heart rate. The input data for time-domain indices such as mHR, SDNN, and RMSSD were fixed at 300 beats, which is the maximum heartbeat in a 3-min recording (Fig. 1). To create the time-domain input data (300-beat data), Raw-1 was placed in the middle, and the rest were filled with the average value of Raw-1. This 300-beat data served to predict the time-domain indices.

**Figure 1.**
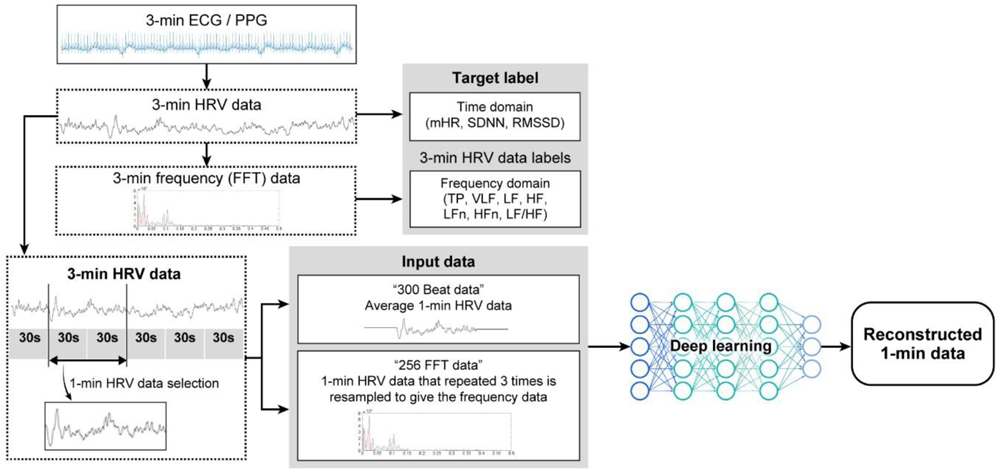
Generation of input data; 3-min HRV data (Raw-3) were generated from each 3-min ECG/PPG. The time and frequency data from Raw-3 served as reference (target label). From Raw-3, a 1-min HRV recording (Raw-1) was selected after 30 s of testing. The data of the time and frequency domains should be uniform in size for training the deep learning models; therefore, the time and frequency domain data were fixed at 300 and 256 beats, respectively. These 300 beats and 256 FFT data served as the input for the deep learning models to predict the outcome, which is the reconstructed 1-min data (Rec-1).

The input data sizes for the frequency domain indices, such as TP, LF, HF, VLF, LFn, HFn, and LF/HF, were fixed at 256 beats. Raw-1 was resampled at 2 Hz and repeated three times. The frequency data from this process (256 FFT data) served as the input to predict the frequency domain indices.

The time and frequency domain labels from Raw-3, 300-beat data, and 256 FFT data were normalized between 0 and 1 to be processed through the deep learning models.

### 2.4 Learning model

From the 34,885 HRV recordings, 20,931 (60%), 10,465 (30%), and 3,489 (10%) were randomly selected for training, validation, and testing, respectively.

A few studies have predicted time dependent indicators using short-term measures; however, there are no reports on measurements that require longer testing time, i.e., LF and VLF. We attempted to measure LFn with a brute force method in 32 empirical models (Supplemental Table 1). Deep Neural Network (DNN) and Convolutional Neural Network (CNN) learning models yielded the best outcomes. We tried other indicators in various types of DNN and CNN models using a brute force method to identify the best learning model for training a particular indicator to find which returned the smallest mean absolute error (MAE) (Supplemental Table 2) [10–13].

Tables 2 and 3 present the learning model used for each indicator. In the case of SDNN, the learning model was CNN2D. The loss function, metric, and optimizer were Huber loss, MAE, and Adam, respectively (Supplemental Figure 1). The learning rate, optimal epoch, and batch size were 1 x 10-5, 14,150, and 5000, respectively.

**Table 2.**
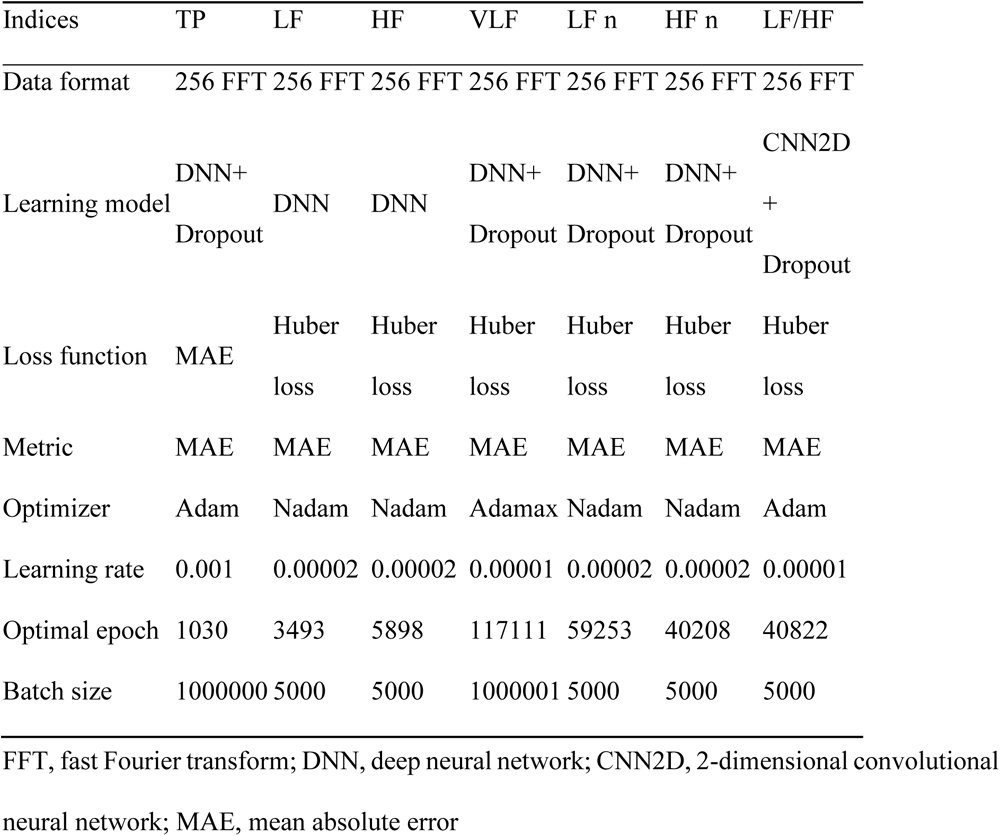
Frequency domain indices and learning models.

**Table 3.** Learning model for the time-domain indices.

| Indices | mHR | SDNN | RMSSD |
| --- | --- | --- | --- |
| Data format | 300-Beat | 300-Beat | 300-Beat |
| Learning model | DNN | CNN2D | CNN1D |
| Loss function | Huber loss | Huber loss | Huber loss |
| Metric | MAE | MAE | MAE |
| Optimizer | Adam | Adam | Adam |
| Learning rate | 0.0001 | 0.00001 | 0.00001 |
| Optimal epoch | 50,873 | 14,150 | 157,584 |
| Batch size | 5,000 | 5,000 | 5,000 |

### 2.5 Statistical processing

The results were analyzed using an independent sample t-test and Pearson’s correlation coefficient. The degrees of consistency between Raw-3 vs. Rec-1 and Raw-3 vs. Raw-1 values were determined using the Bland–Altman plot, and the tolerance limit between the two measured values was defined as average ± 2 × standard deviation (SD). The significance level was set at p < 5 % for all the statistical analyses, and the SPSS19 and Excel technological statistics packages were used.

### 2.6 Institutional review board statement

This study was approved by the review board of the Ajou University Hospital (AJIRB-DEV-MDB-21-258) and was conducted according to the Helsinki Declaration. Patient consent was waived because of the retrospective nature of the study and the analysis used anonymous clinical data.

## 3. Results

### 3.1 Comparison between the data of Raw-3 and Raw-1 vs. Raw-3 and Rec-1

While all the Raw-1 measurements significantly differed (p≤0.05) from those of Raw-3, with the exception of mHR and RMSSD, the measurements of Rec-1 did not show any significant difference in any indices (Table 4). For example, the TP measurements for Raw-3, Rec-1, and Raw-1 of the first subject were 1418.7233, 775.6076, and 282.5084, respectively. For the second subject, the values were 1246.5504, 1148.8865, and 1681.5488, respectively. The Raw-3 and Rec-1 TP measurements of the first 99 subjects were plotted for visual comparison (Fig. 2).

**Figure 2.**
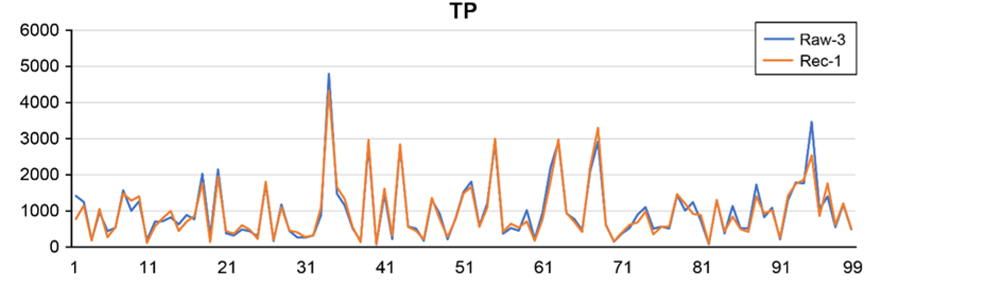
Comparison of TP measurements of Raw-3 and Rec-1. Raw-3 (blue) and Rec-1 (orange) measurements of the first 99 of the 3,489 study subjects are shown. The patterns of the blue and orange lines are similar.

**Table 4.**
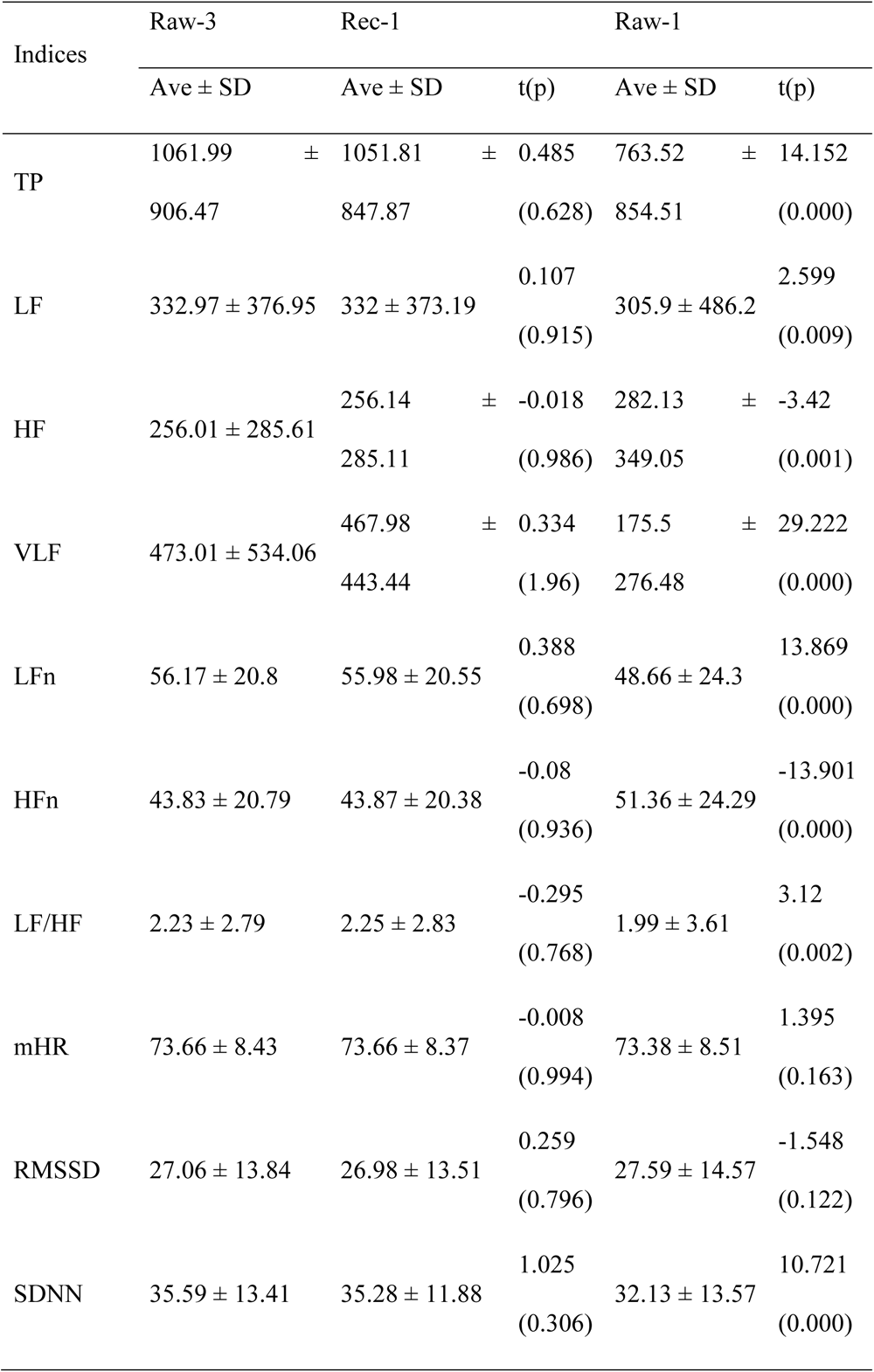
Independent sample t-test of the HRV data.

In most of the 3,489 cases, Rec-1 more closely approximated Raw-3 than Raw-1. The TP of Raw-3, Rec-1, and Raw-1 was 1061.99 ± 906.47, 1051.81 ± 847.87, and 763.52 ± 854.51, respectively, in terms of average ± SD. The TP of Rec-1 (p = 0.628) was not significantly different from that of Raw-3, whereas that of Raw-1 (p = 0.000) was different. The measurements of the other frequency and time-domain indices are presented in Table 4.

### 3.2 Pearson’s correlation between the Raw-3 vs. Rec-1 and Raw-3 vs. Raw-1 indices

The measurements of all the Rec-1 indices showed a good correlation with those of Raw-3. The coefficient of determination (R2) demonstrated that Rec-1 was more strongly correlated with Raw-3 than Raw-1 in the frequency domain indices. The R2 of the time-domain indices of Rec-1 was similar to that of the time-domain indices of Raw-3. The R2 of Raw-3 vs. Rec-1 and of Raw-3 vs. Raw-1 for TP was 0.9307 and 0.4727, LF was 0.9773 and 0.4421, HF was 0.9929 and 0.8147, VLF was 0.6979 and 0.2433, LFn was 0.9553 and 0.5177, HFn was 0.9579 and 0.5167, LF/HF was 0.9099 and 0.4345, mHR was 0.982 and 0.981, RMSSD was 0.9537 and 0.9597, and SDNN was 0.7811 and 0.7681, respectively. Thus, we can conclude that Rec-1 shows a strong correlation with Raw-3.

### 3.3 Prediction error between the indices of Rec-1 and Raw-3

The Bland–Altman plot in Figure 4 shows the difference in each measurement between Rec-1 and Raw-3 for all indices. The x-axis represents Raw-3, and the y-axis represents the difference between Rec-1 and Raw-3. The upper and lower limits of the LoA are expressed as bias ± 1.96 SD, and the number inside the parentheses is the percentage of data within the LoA. For example, for HFn, where Raw-3 was 30.29, its Rec-1 was 36.52; therefore, the prediction error was 6.23, and a blue dot was placed in the coordinate (30.29, 6.23). This plot shows the magnitude of the prediction error. The scattering of points in all indices show that the Rec-1 is in good agreement with the Raw-3.

**Figure 3.**
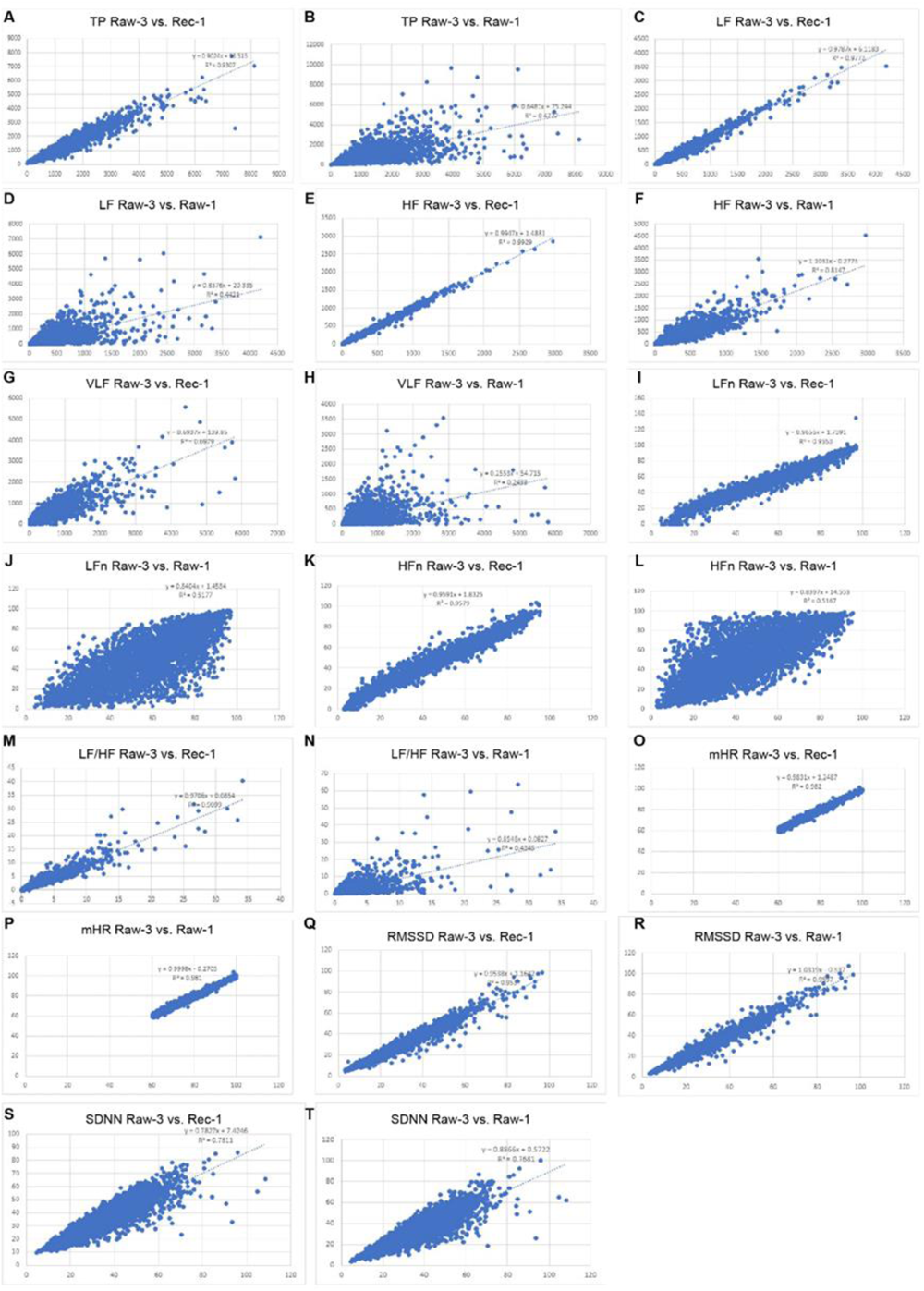
Correlation of indices between Raw-3 vs. Rec-1 and Raw-3 vs. Raw-1. The coefficients of determination (R2) for TP, LF, HF, VLF, LFn, HFn, LF/HF, mHR, RMSSD, and SDNN are shown. Among all frequency domain indices, Rec-1 demonstrated a better correlation with Raw-3.

**Figure 4.**
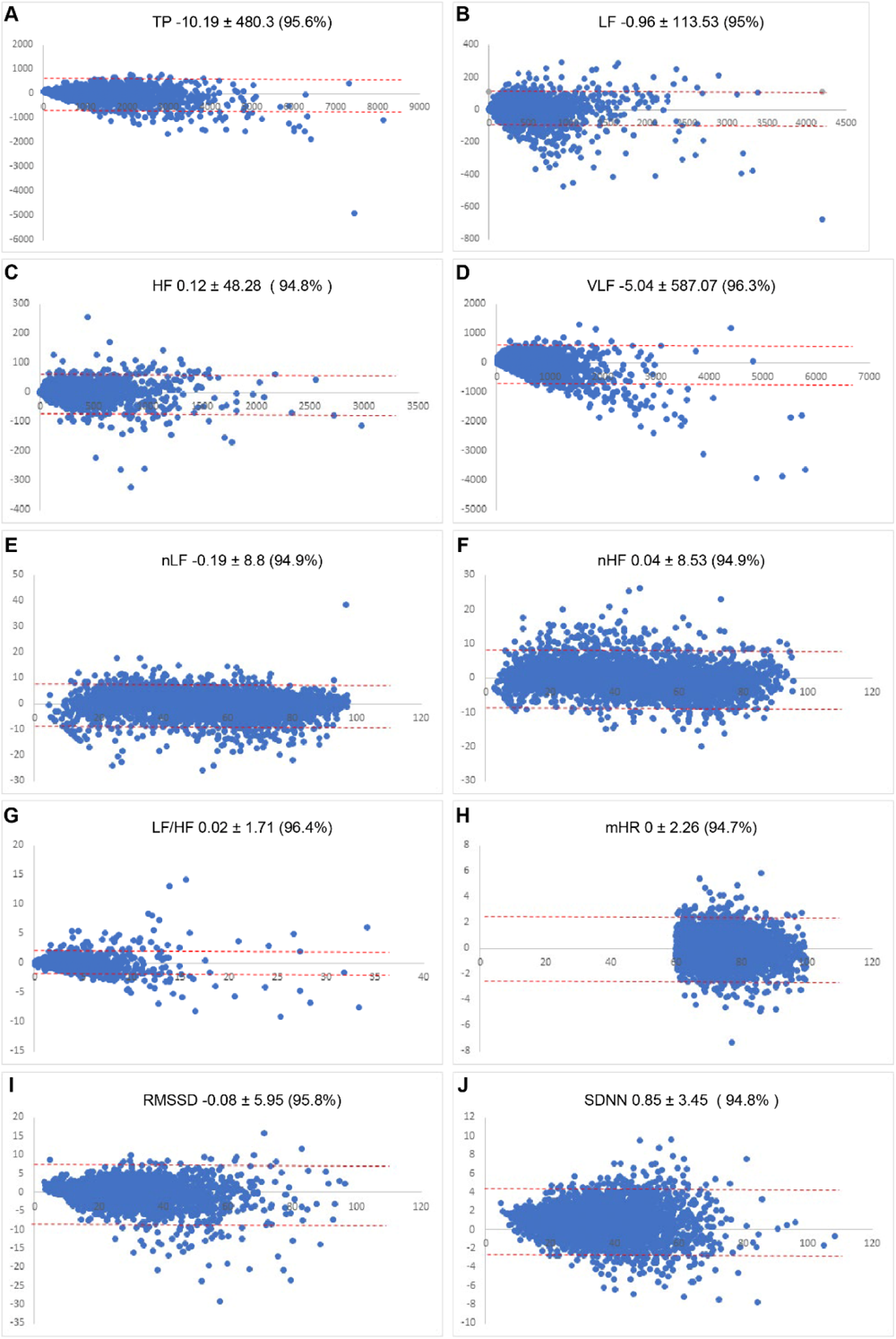
Agreement of Rec-1 and Raw-3. The differences (Raw-3 – Rec-1) in the measurements of all indices are scatter-plotted. The red-dotted lines show 95% confidence interval of the LoA.

## 4. Discussion

Long-term recording (≥ 24 h) reliably assesses lower-frequency indices and is the reference standard for clinical evaluation because of its superior predictive validity [4,14]. Predictive outcomes such as mortality after acute MI and diabetic neuropathy are best assessed with long-term measurements, whereas changes in RR intervals during and immediately after abnormal respiratory events, such as obstructive sleep apnea, can be best assessed with ultrashort-term measurements [15–17].

Despite the benefits of HRV testing in health care, the 5-min recording time is impractically long compared with other clinical tests, such as blood pressure measurement and fasting blood sugar, which can be routinely performed in a primary care center or at home. Interest in shorter measurement times prompted researchers to investigate in ultrashort-term HRV test to assess its reliability against short-term tests [18–20]. Previous studies that tested the minimum testing time showed a high correlation between ultra-and short-term analyses [7–9]. Minimum requirement time for each HRV indices differed, for instance 10 seconds(s) for HR, 20 s for HF, 30 s for RMSSD, and 90 s for LF [8].

Our study challenged the time limit by employing a deep learning algorithm. In this study, the results of independent samples t-tested across all indices showed no significant differences between the groups, including the indices sensitive to measurement time, such as SDNN, VLF, and TP (p < 0.05). The Pearson’s correlation coefficient showed very high correlations, from a minimum of 0.78 (SDNN) to a maximum of 0.99 (HF), with p < 0.000 for both. In the Bland–Altman plot, the difference in most indices fell within the LoA. This indicates that the raw data and predicted values are in good agreement, with minimum tolerable errors.

The coefficient of correlation from previous research that compared 5 and 3-min HRV data was higher for time-sensitive indices such as mHR (0.998 vs. 0.982), SDNN (0.961 vs. 0.781), RMSSD (0.986 vs. 0.953), and VLF (0.866 vs. 0.698) compared with that of our study’s results [2]. However, our results showed better correlations in frequency measures such as TP (0.923 vs. 0.931), HF (0.987 vs. 0.993), and LF (0.955 vs. 0.977).

HRV testing in 1-min allows for a quick assessment of the autonomic nervous system function, which can have various health uses from surveillance to treatment. Myocardial infarction survivors could benefit from frequent HRV screening to stratify future arrhythmic risk, and individuals with chronic heart failure could easily assess disease severity by routinely measuring their HRV at home [21,22]. Patients with psychological conditions such as depression, anxiety, and panic disorder could benefit from HRV biofeedback as adjunctive therapy in inpatient or outpatient settings [23–25].

Despite the findings, the learning process and structural issues associated with deep learning should be considered for further improvement. Larger error values tend to result from greater measurement values. Also, there were cases in which the deviation of some predicted values was greater; however, the error was small for the group average because the learning was based on the absolute error of the group average.

In summary, we employed deep learning methods in HRV analysis for the first time to reduce the recording time to 1-min, and our prediction accuracy was comparable to that of 3-min HRV measurements. Ultrashort-term HRV measurements with deep learning could contribute to the possibility of a nearly real-time health assessment.

## 5. Conclusions

This study demonstrated that a 1-min HRV measurement reconstructed by deep learning can reliably predict 3-min HRV data. Our findings suggest the possibility of assessing the autonomic nervous system function in real time using HRV.

## Data Availability

The data is available upon request to the corresponding author.

## Acknowledgments

We thank Ajou university medical information and media center for the graphic visualization.

